# Development of antibodies to pan-coronavirus spike peptides in convalescent COVID-19 patients

**DOI:** 10.1101/2020.08.20.20178566

**Authors:** Andrii Rabets, Galyna Bila, Roman Grytsko, Markian Samborsky, Yuriy Rebets, Sandor Vari, Quentin Pagneux, Alexandre Barras, Rabah Boukherroub, Sabine Szunerits, Rostyslav Bilyy

## Abstract

Coronaviruses are sharing several protein regions notable the spike protein (S) on their enveloped membrane surface, with the S1 subunit recognizing and binding to the cellular receptor, while the S2 subunit mediates viral and cellular membrane fusion. This similarity opens the question whether infection with one coronavirus will confer resistance to other coronaviruses? Investigating patient serum samples after SARS-CoV-2 infection in cross-reactivity studies of immunogenic peptides from Middle East respiratory syndrome coronavirus (MERS-CoV), we were able to detect the production of antibodies also recognizing MERS virus antigens. The cross-reactive peptide comes from the heptad repeat 2 (HR2) domain of the MERS virus spike protein. Indeed, the peptide of the HR2 domain of the MERS spike protein, previously proven to induce antibodies against MERS-CoV is sharing 74% homology with the corresponding sequence of SARS-CoV-19 virus. Sera samples of 47 convalescent SARS-CoV-2 patients, validated by RT-PCR-negative testes 30 days post-infection, and samples of 40 sera of control patients (not infected with SARS-CoV-2 previously) were used to establish eventual cross-bind reactivity with the MERS peptide antigen. Significantly stronger binding (p< 0.0001) was observed for IgG antibodies in convalescent SARS-CoV-2 patients compared to the control group. If used as an antigen, the peptide of the HR2 domain of the MERS spike protein allows discrimination between post-Covid populations from non-infected ones by the presence of antibodies in blood samples. This suggests that polyclonal antibodies established during SARS-CoV-2 infection has the ability to recognize and probably decrease infectiveness of MERS-CoV infections as well as other coronaviruses. The high homology of the spike protein domain suggests in addition that the opposite effect can also be true: coronaviral infections producing cross-reactive antibodies affective against SARS-CoV-19. The collected data prove in addition that despite the core HR2 region being hidden in the native viral conformation, its exposure during cell entry makes it highly immunogenic. Since inhibitory peptides to this region were previously described, this opens new possibilities in fighting coronaviral infections.

## 1. Introduction

Coronaviruses such as the Middle East respiratory syndrome coronavirus (MERS-CoV), Severe acute respiratory syndrome coronavirus (SARS-CoV-1) and the recently emerged SARS-CoV-2 are sharing several similar protein regions which are involved in the recognition of the host cells. The SARS-CoV-2 genome (30 kb in size) encodes a large, non-structural polyprotein (ORF1a/b) that is further proteolytically cleaved to generate 15/16 proteins, 4 structural proteins and 5 accessory proteins (ORF3a, ORF6, ORF7, ORF8 and ORF9). The four structural proteins consist of the spike (S) surface glycoprotein, the membrane (M) protein, the envelope (E) protein and the nucleocapsid (N) protein, which are essential for SARS-CoV-2 assembly and infection. The MERS-CoV genome structure is encoding 10 proteins; two replicase polyproteins (open reading frames [ORFs] 1ab and 1a), three structural proteins (E, N, and M), a surface (spike) glycoprotein (S), and five nonstructural proteins (ORFs 3, 4a, 4b, and 5) [1,2].

The spike surface glycoprotein (S) plays a key role in mediating virus attachment and fusion and are indeed present in all human infecting coronaviruses. They can be cleaved by host proteases into an N-terminal S1 subunit and a membrane-bound C-terminal S2 region. In order to engage a host receptor, the receptor-binding domain (RBD) of the S1 subunit undergoes conformational movements, which transiently hide or expose the determinants of receptor binding [3,4]. The Heptad Repeat 1 (HR1) region in S2 subunits forms a homotrimeric structure, exposing 3 highly conserved hydrophobic grooves on the surface resulting in binding of 3 Heptad Repeat 2 (HR2) regions and formation of six-helix bundle structure (6-HB). 6-HB is responsible for close approximation of viral and host membranes and their subsequent merging. Binding of HR1 and HR2 domains results in the six-helix bundle needed for merging with host cell membrane. Thus parts of the HR2 domain specifically binding HR1 can be considered as ideal coronaviral inhibitor strategy preventing cellular entry [5]. Further optimization of peptide sequence resulted in pan-coronaviral inhibitors, like EK1 [3] able to inhibit SARS-CoV-19 pseudovirus infection [6].

The structure of HR2 is poorly resolved during crystallographic assessment due to high level of flexibility [7]. Unlike highly mutable receptor-binding domain, the HR1 and HR2 domains are highly conservative between coronaviruses, so form a perfect target for viral neutralization and generation of immunity that latter can be used for viral testing. Since HR2 and HR1 domains are merged and surface exposed after S protein cleavage we expected them to be highly immunogenic.

This similarity can result in the development of cross-reactive antibodies and protection against other coronaviruses, in case of being infected by another virus species. In this work we would like to establish if SARS-CoV-2 results in production of antibodies, that are also recognizing MERS virus antigens.

## 2. Results

### 2.1. Conformation agreement between S1 MERS peptide with the genome of SARS-CoV-2

The complete crystal structure of the HR2 domain of SARS-CoV-2 remains currently unavailable due to it conformation changes and problems in stabilization [4] (**Figure 1a**). Using SEQATOMS algorithm [8] the most complete structure of the corresponding region containing defined atomic coordinates was identified to be the one proposed by Walls et al [9], namely a model for HR1 HR2 rearrangements and unfolding accompanying viral entry into host cells for other coronaviruses. This structure clearly demonstrates exposure of the HR2 domains upon cellular binding in trimeric (**Figure 1b**) and monomeric form (**Figure 1c**). To evaluate the cross reactivity we selected the HR2-specific peptide of the spike protein of MERS-CoV reported to possessed high immunogenic potential [10–12]. Indeed, ongoing studies showe that it can be also successfully used to raise MERS-recognizing antibodies in the presence of neutrophil extracellular traps(NET) forming nanoadjuvants [13]. The selected HR2 peptide of the spike protein of MERS virus (depicted yellow, **Figure 1d** using PDB deposited crystal structure of 4NJL_A, [14]) shares significant similarities in 3D structure between (the only) known crystal structure of unfolded HR2 domain (**Figure 1c**) and with pan-coronaviral inhibitor peptide EK1, using crystal structure 5ZVK_a [3] (**Figure 1e**). Protein BLAST analysis reveales 46% identity and 76% similarity in aminoacid sequence of the MERS peptide with the corresponding peptide of the SARS-CoV-19 spike protein (sequence ID QKJ68605.1) (**Figure 1f**).

**Figure 1.**
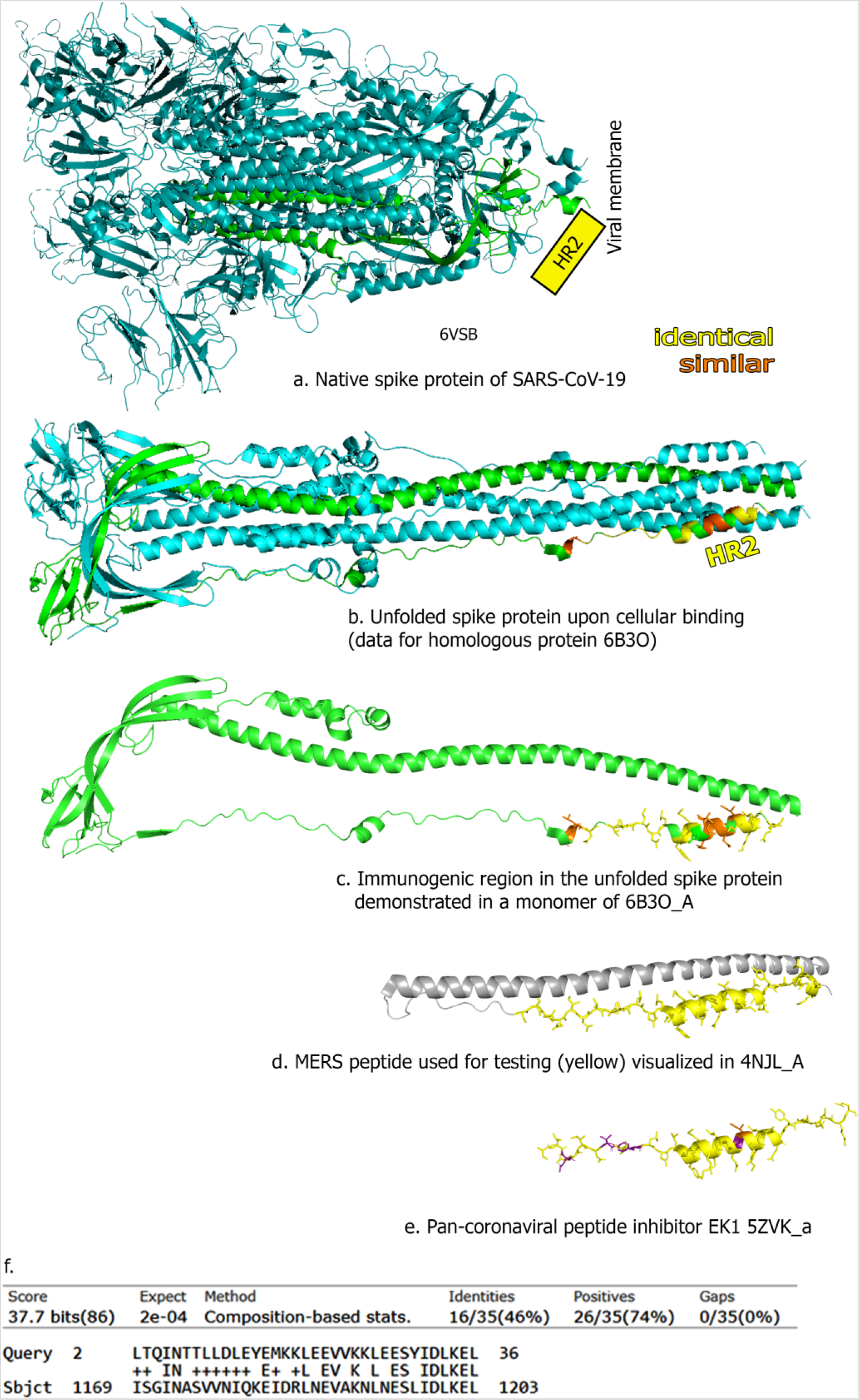
Similarity of HR2 regions between different coronaviruses. (a) Image of Spike protein (S1) of SARS-CoV-2. (b) S1 upon host cell interaction: conformational changes in trimer are occurring, exposing previously hidden HR2 domain regions [4], (c) – monomeric part of (b), exposing domain with structural similarity (yellow – identical, orange – similar amino acids) towards corresponding MERS peptide, depicted on (d) and pan. (f). Protein BLAST analysis of used MERS protein is showing similarity towards the sequence in the genome of SARS-CoV-19. Sequence ID: QKJ68605.1. Crystal structure of HR2 domain for SARS-CoV-19 is currently not available, thus it is represented as rectangle in (a) based on last connected coordinates available in 6VSB structure.

### 2.2. Antibody cross-reactivity towards coronaviral HR2 domains

Having established the structural similarity between the S1 MERS peptide with the genome of SARS-CoV-2, sera of convalescent SARS-CoV-2 infected patient, who have never suffered from MERS-CoV infection before, have been collected and tested for the presence of antibodies. The S1 MERS-CoV specific peptide, NH_2-_CCTTTTTTSLTQINTTLLDLEYEMKKLEEVVKKLEESYIDLKEL-COOH, which we previously successfully used to raise anti-MERS antibodies while testing novel NET-stimulating adjuvants [13], was immobilized on ELISA plates and incubated with sera samples. As can be seen from **Figure 2a**, stronger binding of IgG from sera of SARS-CoV-2 convalescent patients is observed when compared to sera of patients without previous SARAS-CoV-2 infection (p < 0.0001, n = 87). Minor differences were detected for IgM binding (p = 0.016) and no difference was detected for IgA or IgE antibody subclasses (supplementary information **Figure S1**).

**Figure 2.**
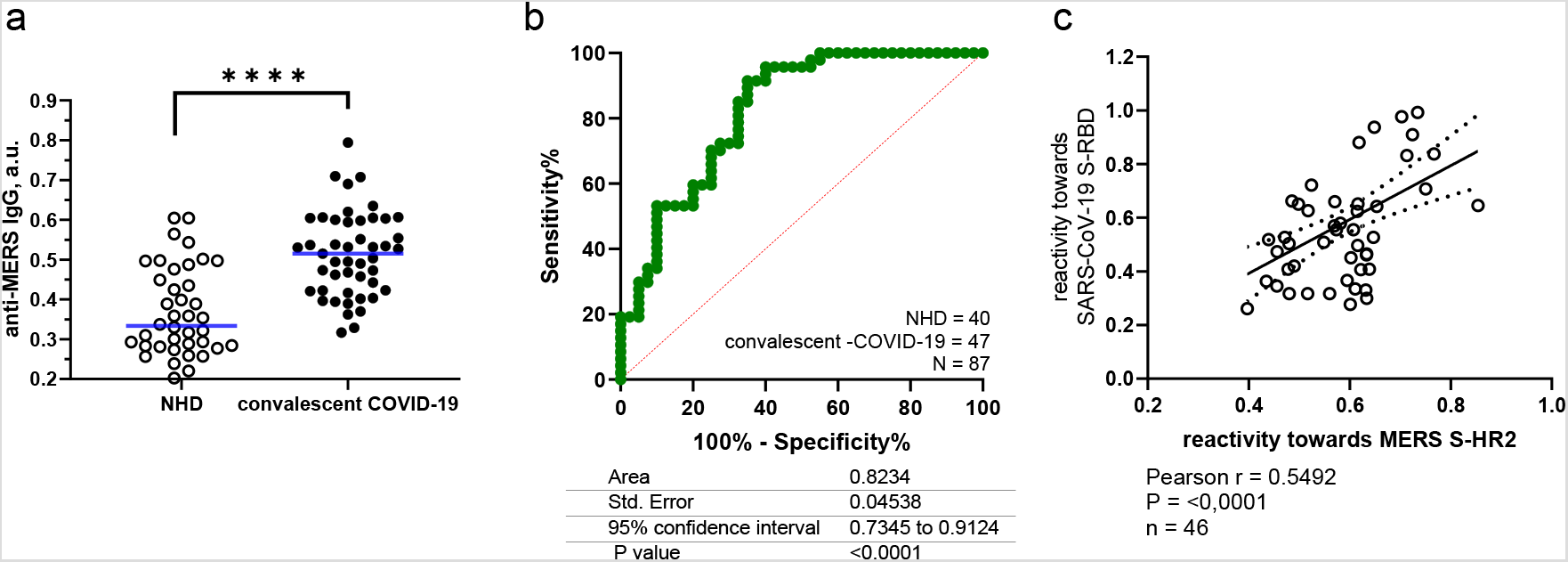
Antibody cross-reactivity between coronaviruses. serum of convalescent COVID-19 patients possess antibodies recognizing MERS-specific peptide of HR2 spike protein domain. a. difference between IgG levels recognizing peptide of HR2 domain in S protein of MERS in convalescent SARS-CoV-2 patients and non-infected healthy donors. b – ROC curve for discrimination of “anti-coronaviral” IgG antibodies using indicated MERS peptide as antigen. c – IgG reactivity in convalescent plasma of SARS-CoV-2 patients towards HR2 domain in S protein of MERS versus RBD part of S protein of SARS-CoV-19.

Using this S1 MERS-CoV specific peptide, discrimination of persons that have suffered from SARS-CoV-2 infection and those who were not in contact with the virus resulted in a predictive value (area under ROC curve) equal to 0.823 (**Figure 2b**), with a specificity and sensitivity of ∼60% (95% confidence). SARS-CoV-2 infections results in the generation of antibodies with significantly strong cross-reactive towards a MERS specific peptide with 76% homology. Highly conservative region of the exposed domain suggest that the opposite can be true – coronaviral disease can result in some antibodies able to recognize SARS-CoV-19 epitops circulating in the blood.

To determine whether the strong binding with S1 peptide is correlated with higher amount of anti-SARS-CoV-19 antibodies we used recombinant RBD protein immobilization on ELISA plates to incubate with sera samples to evaluate the amount of formed IgG type antibodies. As can be seed from **Figure 2c**, the sera samples having shown stronger binding of IgG antibodies with anti-HR2 MERS spike protein also contained higher IgG reactivity towards anti-RBD spike protein of SARS-CoV-19. The Pearsons correlation between the two parameters was 0.5492, p< 0.0001. This suggests stronger humoral responses towards one of the virus will be associated with the intensity of the immune response towards other coronaviruses.

## 3. Discussion

Cross-reactivity between coronaviruses has become a critical question, since it brings new promises against COVID-19 protection. On the other hand, this cross-reactivity can be negative, since available cross-reactivity towards coronaviruses will make coronaviruses not the best choice for vectors in vaccines, especially taking into account recent data on broad immune cross-reactivity [15]. Indeed, it was reported that epitope pools detect CD4+ and CD8+ T cells in 100% and 70% of convalescent COVID patients respectively, recognizing S and M proteins, with at least eight SARS-CoV-2 ORFs targeted. T cell reactivity to SARS-CoV-2 epitopes is also detected in non-exposed individuals [6]. In SARS-CoV-2 patients S-reactive CD4+ T cells equally target N-terminal and C-terminal parts of the spike protein, whereas in healthy donors S-reactive CD4+ T cells react almost exclusively to the C-terminal part. This part is characterized by a higher homology to spike glycoprotein of human endemic “common cold” coronaviruses, and contains the S2 subunit of S with the cytoplasmic peptide (CP), the fusion peptide (FP), and the transmembrane domain (TM) but not the receptor-binding domain (RBD). S-reactive CD4+ T cells from SARS-CoV-2 patients are further distinct to those from healthy donors as they co-expressed higher levels of CD38 and HLA-DR, indicating their recent *in vivo* activation [7]. Potential preexisting cross-reactive T cell immunity to SARS-CoV-2 has broad implications, as it could explain aspects of differential SARS-CoV-2 clinical outcomes, influence epidemiological models of herd immunity, or affect the performance of SARS-CoV-2 candidate vaccines. Pre-existing memory CD4+ T cells that are cross-reactive with comparable affinity to SARS-CoV-2 and the common cold coronaviruses HCoV-OC43, HCoV-229E, HCoV-NL63, or HCoV-HKU1. Thus, variegated T cell memory to coronaviruses that cause the common cold may underline at least some of the extensive heterogeneity observed in COVID-19 disease [8].

Based on these and our data one can assume that the C-terminal part of the spike protein is immunogenic due to exposure upon merging with host cells. Its conservative nature provides the background for the development of cross-coronaviral immune responses both cellular, and demonstrated here, by a IgG type humoral immune response.

## 4. Material and methods

### Patient cohorts

All analyses of human materials were performed in accordance with the institutional guidelines and with the approval of the Ethics Committees of Danylo Halytsky Lviv National Medical University (DH LNMU No.5/2017–02–23). The current study involved 47 samples from SARS-CoV-2 infected patients treated in the Lviv Regional Clinical Infection Hospital of Infectious Diseases / Department of Infectious Diseases, Danylo Halytsky Lviv National Medical University (Lviv, Ukraine) during April-May 2020. Written informed consent was obtained from every person before blood collection. All patients were PCR-positive upon hospitalization. Sera collection took place at least 3 weeks after the appearance of clinical symptoms, when person were recovered. Recovery was assessed by a) disappearance of clinical symptoms, b) two consecutive negative PCR tests made within 2 days difference. Group of normal healthy volunteers, who have donated blood between June and November 2019 (pre-COVID-19) served as controls. Informed written consent for blood withdrawal was obtained from each patient and NHD.

### ELISA tests

Sera samples from convalescent COVID patients and NHDs were frozen at –20°.

For testing anti-MERS response immunosorbent NUNC maxisorp plates (Thermo Scientific, Waltham, USA) were coated with NH_2-_CCTTTTTTSLTQINTTLLDLEYEMKKLEEVVKKLEESYIDLKEL-COOH peptide (50 μL of a 4 μg/mL solution) in carbonate-bicarbonate buffer (100 mM, pH 9.6). For testing anti SARS-CoV-19 response immunosorbent NUNC maxisorp plates (Thermo Scientific, Waltham, USA) were coated with recombinant 194 a.a. protein corresponding to RBD domain of spice protein of SARS-CoV-19, (Exploregen LLC, UA). All serum samples were diluted 1:1000 in carbonate-bicarbonate buffer and incubated at 37°C for 1 h, after that the plates were washed again.

Goat anti-human IgG (H+L)-horseradish peroxidase (HRP) (109–035–003, Jackson ImmunoResearch) was diluted in washing buffer (1:25000), added to the plates and incubated at room temperature for 1 h. After the corresponding washings, the assay was developed with 3,3’,5,5’-tetramethybezidine (TMB) containing excess of H_2_O_2_ as a substrate (50 µL per well). The reaction was stopped with 50 µl/well of sulfuric acid (1 M). The absorbance was read at 450 nm/600nm using a Perkin Elmer BioAssay reader HST700 (Waltham, USA).

### Bioinformatics

The protein homology searches were done using blast (NCBI) and PDB databases. In order to include the regions with resolved structures in our searches we had used SEQATOMS (http://www.bioinformatics.nl/tools/seqatoms/) [8]. Protein structures were visualized using PyMOL (https://pymol.org/). Multiple sequence alignments were done using CLUTALW [16]

### Data analysis

ELISA testing was performed in duplicate using 2 technical replicates for each analysis (coefficient of variation [CV] always < 3%). The data were normalized between plates using positive controls and corrected for background signal of secondary antibodies, then the mean values were calculated and are shown on the graphs. For comparisons between two groups, the Mann–Whitney U-test for numerical variables was used. A receiver operating characteristic (ROC) curve was generated. The area under the ROC (AUROC) was calculated to estimate the specificity, sensitivity and usefulness of the binding assays. All analyses were performed using Excel 2016 (Microsoft Corp., Redmond, WA, USA) and Prism 8.2 (GraphPad, San Diego, USA) software. A p-value of ≤0.05 was considered statistically significant. Four levels of significance are depicted in the figures by asterisks: *– p< 0.05; **– p< 0.01; ***– p< 0.001; ***– p< 0.0001.

## Data Availability

Data are available upon justified request.

## Acknowledgements

We thank Prof. Jean Dubuission and Dr. Karin Seron, University of Lille, CNRS, INSERM, CHU Lille, Institut Pasteur de Lille, U1019 - UMR 8204 - CIIL - Center for Infection and Immunity of Lille, F-59000 Lille, France for providing immunogeneic MERS peptides. Financial support from the Cedars Sinai Medical Center’s International Research and Innovation in Medicine Program, the Association for Regional Cooperation in the Fields of Health, Science and Technology (RECOOP HST Association) RCSS grant 020, and BMYSRG 015; Grant of Ministry of Healthcare of Ukraine 0119U101338; Volkswagen-Stiftung grant No 97744. This project has received funding from the European Union’s Horizon 2020 research and innovation programme under grant agreements No 861878 and 872331. Financial support by ANR project “nanoMERS” (ANR-18-CE09–0021) is acknowledged

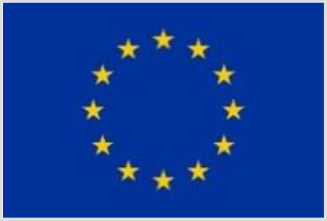

---

**Figure S1.**
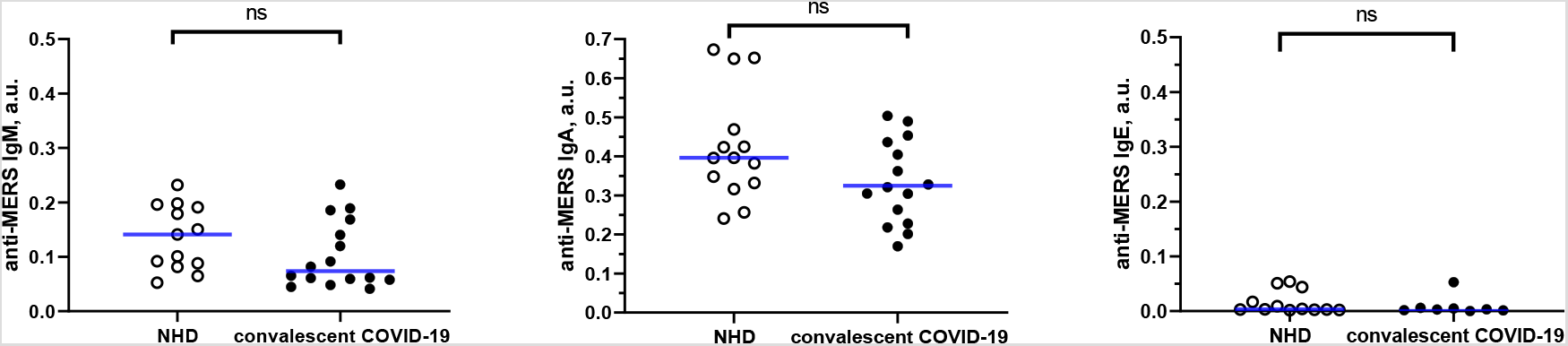
Antibody classes binding peptide of HR2 spike protein of MERS virus in convalescent COVID-19 sera, from left to right: binding of IgM, IgA and IgE immunoglobulins.

